# Determinants of an Unintended Pregnancy among Women Attending Ante Natal Care at Health Facilities in Lemi Kura Sub City, Addis Ababa, Ethiopia: Institution based Case-Control Study

**DOI:** 10.1101/2025.06.26.25330390

**Authors:** Amideyesus Adinaw lopiso, Teketel Tumebo, Sisay Tanie, Kaleab Tadesse worku, Nebiyu Dereje, Tamane Achamo, Andamlak Gizaw Alamdo

## Abstract

**Background:** Unintended pregnancy refers to a pregnancy that is either unplanned at the time of conception or not wanted at all. It carries significant social, emotional, and health consequences for women, their children, families, and communities. In Ethiopia, approximately one-third of married women report that their pregnancies are unintended. Despite the considerable burden of unintended pregnancies in the country, there remains limited evidence on the factors contributing to this issue, particularly at Lemi Kura sub city. This study, therefore, aimed to identify the determinants of unintended pregnancy among women attending antenatal care services in health facilities within Lemi Kura sub-city, Addis Ababa, Ethiopia.

**Methods:** We conducted a facility-based case-control study in Lemi Kura sub-city, Addis Ababa, Ethiopia, from January 3 to February 18, 2022. Cluster random sampling technique was used to select 103 cases (women with unintended pregnancies attending antenatal care unit) and 197 controls (women with planned pregnancies attending antenatal care unit). Data were collected through interviewer-administered, pretested questionnaires. The sample size was calculated using Epi-Info version 7. Data analysis was carried out using SPSS version 20, applying multivariable logistic regression to identify determinants of unintended pregnancy. Results were reported as adjusted odds ratios (AOR) with 95% confidence intervals (CI), and statistical significance was declared at *p* < 0.05.

**Results:** The Muslim women (AOR: 3.33, 95% CI: 1.21–9.15); Protestant women (AOR: 3.06, 95% CI: 1.07–8.75); reduced women’s autonomy (AOR: 16.58, 95% CI: 2.40–114.15); residing 4 km or more from the antenatal care facility (AOR: 2.70, 95% CI: 1.07–8.75); having two or more living children(parity) (AOR: 5.37, 95% CI: 1.60–18.15); desiring two or fewer children(Ideal) (AOR: 3.63, 95% CI: 1.50–9.01); and having multiple sexual partners (AOR: 4.25, 95% CI: 1.38– 30.38) were significant predictors of unintended pregnancy.

**Conclusion:** Women with Low knowledge about FPMs, unmarried, Protestant and Muslim religious, low autonomy, multiparous, used natural FPM, health service inaccessibility and having multiple sexual partners were primary predicators of the unintended pregnancy. For intervention, Engaging religious leaders and focusing on multiparous with empowering women is essential.

## Introduction

Unintended pregnancy (UP) is a significant public health concern worldwide, referring to pregnancies that are either mistimed or unwanted at the time of conception. A mistimed pregnancy occurs earlier than desired, typically defined as occurring two or more years before a woman intended to conceive (<u>1</u>), whereas an unwanted pregnancy is one that is not desired at any point in the future (<u>2</u>). Unintended pregnancies have diverse outcomes, including abortion, miscarriage, or unplanned birth, with profound implications for maternal and child health.

Unintended pregnancies contribute substantially to maternal morbidity and mortality, particularly in low-resource settings where access to maternal healthcare services is limited. Pregnancies occurring too early, too late, or too frequently increase the risk of complications during pregnancy, labor, delivery, and the postpartum period (<u>3</u>). Women experiencing unintended pregnancies face significant health risks, economic burdens, and social consequences, including disruptions in education and employment opportunities (<u>2</u>).

The burden of unintended pregnancy is disproportionately higher in developing countries compared to developed nations (<u>4</u>). In 2010, an estimated 80 million unintended pregnancies occurred globally, with approximately 75 million occurring in low-income regions (<u>3</u>). Despite international efforts to address the issue, unintended pregnancy rates remain high, particularly in sub-Saharan Africa (<u>5</u>). Various socio-demographic factors, such as low socioeconomic status, limited access to and inconsistent use of contraceptives, early or late reproductive age, and lower educational attainment, have been identified as key contributors to unintended pregnancies (<u>6</u>).

A major consequence of unintended pregnancy is unsafe abortion, which is associated with serious health complications, including infertility, severe infections, and maternal death (<u>7</u>). A study conducted by the Guttmacher Institute in Ethiopia reported that 101 unintended pregnancies occurred per 1,000 women aged 15-44, with 42% of all pregnancies classified as unintended. In Addis Ababa, the prevalence was even higher, with an estimated 49 unintended pregnancies per 1,000 women (<u>8</u>).

Globally, unintended pregnancies account for a substantial proportion of unsafe abortions. Of the estimated 210 million pregnancies occurring annually, approximately 38% are unintended, and 22% result in abortion (<u>9</u>). Alarmingly, 40% of these abortions are performed on women younger than 25 years of age, with approximately 68,000 maternal deaths occurring each year due to complications from unsafe abortion procedures (<u>10</u>). The burden is particularly severe in developing countries, where maternal and neonatal mortality rates remain disproportionately high. In sub-Saharan Africa, maternal mortality occurs in approximately 1 in 160 pregnancies, underscoring the urgent need for targeted interventions (<u>11</u>).

Several studies have identified key determinants of unintended pregnancy, including parity, age at marriage, media exposure, religious beliefs, and awareness of family planning methods (<u>3</u>). The 2016 Ethiopian Demographic and Health Survey (EDHS) reported that 17% of women described their last pregnancy as mistimed, while 8% categorized it as unwanted (<u>12</u>). Despite the availability of family planning services, unintended pregnancy rates have continued to rise in Ethiopia, exacerbating maternal and child health challenges (<u>3</u>).

Women’s autonomy and accessibility to family planning services play a critical role in the prevention of unintended pregnancies. Limited decision-making power, coupled with gaps in contraceptive availability and use, increases women’s vulnerability to unintended pregnancies (<u>13</u>). In Ethiopia, many women resort to unsafe abortion practices due to unplanned pregnancies, resulting in a range of negative health and social outcomes, including increased risk of maternal mortality, reproductive health complications, economic strain, and psychosocial distress (<u>14</u>).

Lemi Kura sub-city in Addis Ababa, established in 2020, has limited research on the determinants of unintended pregnancy among women attending antenatal care (ANC) services. Given the absence of documented studies in this newly founded sub-city, a case-control study is warranted to assess the key determinants of unintended pregnancy in this population. Understanding these factors will provide valuable evidence for policymakers and healthcare providers, facilitating the development of targeted strategies to reduce unintended pregnancies and improve reproductive health outcomes.

This study aims to identify the determinants of unintended pregnancy among women attending ANC services at Lemi Kura health facilities. The findings will inform evidence-based policymaking, enhance family planning program implementation, and serve as a foundation for future research on reproductive health in Ethiopia.

## Methods and Materials

### 2.1 Study design and setting

An institution-based unmatched case-control study was conducted from January 3,2022 to February 18, 2022, in Lemi Kura Sub-city, Addis Ababa, Ethiopia. Lemi Kura is the newest of the city’s eleven sub-cities, established in 2020, and is bordered by Bole and Yeka sub-cities of Addis Ababa city administration as well as the Oromia Region. In 2022, the sub-city had an estimated population of 344,944, with 165,573 males and 179,371 females. Administratively, it is divided into 10 woredas. According to the 2021 annual report from the sub-city health department, there are 318 health facilities including 126 health service providing clinics and 192 pharmacies (10 public and 308 private). Reproductive health services, including antenatal care (ANC), are offered by 14 health institutions including 4 hospitals, 1 specialty center and 9 health centers, serving an estimated 1,799 ANC visits per month. The study was carried out at one private hospital (ICMC) and three public facilities: Abebech Gobena Hospital, Woreda 13 Health Center, and Hidasse Health Center.

### 2.2 Population and sampling

We conducted an unmatched case-control study among pregnant women attending antenatal care (ANC) at four randomly selected health facilities (three government-run, one private) in Lemi Kura Sub-city, Addis Ababa (January 3-February 18, 2022). Cases were women with unintended pregnancies (unwanted or mistimed by ≤ 2 years), while controls had planned pregnancies (1:2 ratio). For every unintended pregnancy case, two consecutive controls with planned pregnancies were included.

The sample size was calculated using Epi Info 7 (two-population proportion formula) with 95% confidence level, 80% power, and key parameters from prior research (32.1% vs. 16.9% age ≤24 among cases/controls) (<u>18</u>). With a 10% non-response buffer, we enrolled 300 participants (103 cases, 197 controls), proportionally allocated across sites: Hidasse Health Center (n=91), Woreda 13 Health Center (n=95), Abebech Gobena Hospital (n=100), and ICMC Hospital (n=14) (Fig. 1). Women who were critically ill or unable to participate were excluded. The timing of data collection and proportion of cases to controls were maintained in accordance with the allocated sample size.

**Figure 1:**
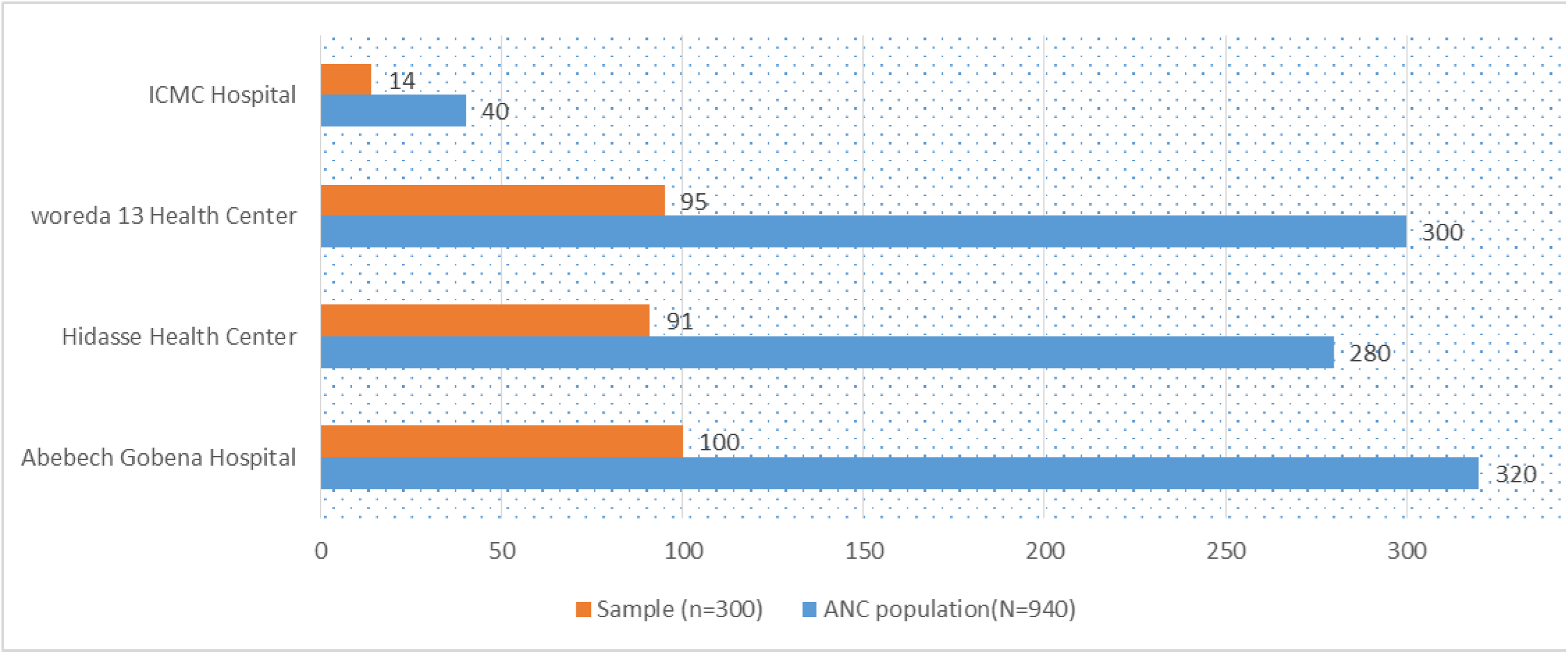
proportional sample size allocation based on average monthly ANC attendance across study sites, Lemi Kura Subcity, Addis Ababa, Ethiopia, 2022.

### 2.3 Data collection tool and measurement

We collected data using a pretested, structured questionnaire adapted from validated instruments in published literature [<u>31</u>,<u>32</u>,<u>33</u>,<u>36</u> & <u>37</u>]. The tool assessed: Socio-demographic characteristics, Obstetric and reproductive health history, Healthcare access and utilization, Behavioral factors, Pregnancy intention (to classify cases/controls).

The questionnaire was developed in English, professionally translated into Amharic and Afan Oromo by bilingual experts, and back-translated to ensure conceptual equivalence.

Well trained personnel conducted face-to-face interviews at the selected health facilities, with a professionally trained eight midwives serving as data collectors supervised by two field supervisors and the principal investigator. To confirm pregnancy intention classification (not routinely documented in medical records), antenatal care providers used indirect questioning techniques, with responses systematically coded. We further validated our instrument through pretesting on 5% of the sample at a non-study health facility.

### 2.4 Data Quality Control

To ensure data quality, we employed standardized questionnaires derived from validated scales and relevant literatures. The instrument was initially developed in English, then professionally translated into Amharic and Afan Oromo by certified bilingual experts, followed by back-translation to verify accuracy. Prior to data collection, we conducted a pretest involving 5% of the calculated sample size (8 cases and 8 controls) at non-selected health facilities within the study area, refining the questionnaire based on these findings.

Eight trained data collectors and two supervisors participated in a one-day intensive training covering study objectives and data collection protocols. To enhance measurement validity, we generated and evaluated composite index scores before conducting statistical analyses, ensuring robust estimation of key study parameters.

### 2.5 Data Processing and Analysis

Data were coded, checked for completeness, and entered into Epi Info 7 before export to SPSS 20 for analysis. Descriptive statistics summarized both cases and control participant characteristics. Bivariate logistic regression identified candidate variables (p < 0.25) for multivariable analysis, where a backward stepwise (likelihood ratio) approach was used to determine significant predictors of unintended pregnancy. Model adequacy was confirmed via Hosmer-Lemeshow goodness-of-fit test (p > 0.05). Associations were reported as adjusted odds ratios (AOR) with 95% confidence intervals (CI), with statistical significance set at p < 0.05.

### 2.6 Operational definition

**Unintended pregnancy (UP):** is unwanted (pregnancy that is not desired now, later or any time in the future) pregnancy or mistimed (pregnancies that are desired either later or sooner) pregnancy.

## Results

### 3.1 Socio demographic characteristics of Respondents

A total of 300 pregnant women (103 cases, 197 controls) attending antenatal care (ANC) were included in the study (100% response rate). They ranged in age from 16 to 43 years (mean ± SD: 27.10 ± 4.73 years), with cases slightly older than controls (mean 27.61 vs 26.84 years, respectively). The majority (91% overall) were married (81.6% of cases and 85.9% of controls) and 89% cohabited with their partners, while the remainder (9% overall; 18.4% of cases and 4.0% of controls) were single, divorced, or widowed. The religious composition was predominantly Orthodox Christian (61.3% overall; 50.0% of cases and 67.0% of controls), with Muslims accounting for 20.3% and Protestants 18.4%. Ethnically, participants were mostly Amhara (45.3%) or Oromo (21.0%). In terms of residence, 62.2% lived in Lemi Kura sub-city of Addis Ababa, 23.0% in other sub-cities of Addis Ababa, and 14.0% in Oromia region. Regarding occupation, 36.0% were housewives, 27.0% were government employees, and 13.9% worked in the private or NGO sector. The mean monthly household income was 10,057.73 ± 7,130.57 birr (10,251 birr among cases vs 9,828.44 birr among controls). Educational attainment was evenly split, with 17% were illiterates and under primary, 28.3% were primary education and 54.6% were secondary and higher education completed. On average, participants lived 2.51 ± 1.62 km from the nearest health facility and 4.28 ± 3.17 km from the current ANC following site <u>(Table 1).</u>

**Table 1:** Socio-demographic characteristics of participants in Lemi kura Sub City, Addis Ababa, Ethiopia, 2022 (n = 300).

| Variables | Categories | Controls(%) | Cases(%) | Total(N) | % | P-value |
| --- | --- | --- | --- | --- | --- | --- |
| Age | 16 to 24 | 55(27.9) | 27(26.2) | 82 | 27.3 | 0.002 |
|  | 25 to 34 | 133(67.5) | 66(64.1) | 199 | 66.3 | 0.969 |
|  | 35 to 43 | 9(4.6) | 10(9.7) | 19 | 6.3 | 0.113 |
| Marital Status | single | 4(2) | 13(12.6) | 17 | 5.7 | 0.039 |
|  | married | 189(95.9) | 84(81.6) | 273 | 91 | 0.001 |
|  | widowed and divorced | 4(2) | 6(5.8) | 10 | 3.3% | 0.370 |
| The dwelling of the participant | same house | 184(93.9) | 82(79.6) | 266 | 89 | 0 |
|  | different place | 12(6.1) | 21(20.4) | 33 | 11 | 0 |
| Religion | orthodox | 132(67) | 52(50.5) | 184 | 61.3 | 0 |
|  | Muslim | 34(17.3) | 26(25.4) | 60 | 20.3 | 0.031 |
|  | Protestant | 31(15.8) | 25(23.3) | 55 | 18.4 | 0.023 |
| Ethnicity | Amhara | 97(49.2) | 39(37.9) | 236 | 45.3 | 0.059 |
|  | Oromo | 36(18.3) | 27(26.2) | 63 | 21 | 0.939 |
|  | Gurage | 16(8.1) | 6(5.8) | 22 | 7.3 | 0.219 |
|  | Tigre | 11(5.6) | 4(3.9) | 15 | 5 | 0.274 |
|  | others | 37(18.8) | 27(26.2) | 64 | 21.3 | 0.213 |
| Occupational status | Housewife | 74(37.9) | 36(35) | 110 | 36.9 | 0.869 |
|  | G.Employee | 53(27.2) | 28(27.2) | 81 | 27.2 | 0.963 |
|  | Merchant | 21(10.8) | 13(12.6) | 34 | 11.4 | 0.714 |
|  | Daily laborer | 20(10.3) | 12(11.7) | 32 | 10.7 | 0.767 |
|  | others | 27(13.9) | 14(5.8) | 41 | 13.4 | 0.046 |
| Educational status | Illiterate | 8(4.1) | 10(9.7) | 18 | 6 | 0.21 |
|  | Read and write | 21(10.7) | 12(11.7) | 33 | 11 | 0.775 |
|  | primary | 62(31.5) | 23(22.3) | 85 | 28.3 | 0.099 |
|  | grade 9-12 | 58(29.4) | 27(26.2) | 85 | 28.3 | 0.318 |
|  | college | 48(24.4) | 31(30.1) | 79 | 26.3 | 0.058 |
| average income of the household | <3000 | 19(9.7) | 10(9.9) | 29 | 9.8 | 0.748 |
|  | 3001 to 10,000 | 108(55.4) | 60(59.4) | 168 | 56.8 | 0.464 |
|  | >10,000 | 68(34.9) | 31(30.7) | 99 | 33.4 | 0 |
| Distance from ANC facility | <4km | 135(71.8) | 63(61.2) | 203 | 10.7 | 0 |
|  | >=4km | 55(28.2) | 40(38.8) | 95 | 31.9 | 0.062 |

### 3.2 Obstetric and Reproductive Characteristics of Respondents

The majority of respondents in both groups reported having information about family planning methods, with 97.5% of controls and 96.1% of cases indicating awareness. Health workers were the primary source of information for 52.3% of controls and 58% of cases, followed by colleagues and others (26.7% of controls vs. 27% of cases), and media such as TV and radio (19% and 13%, control and case respectively).

Regarding beliefs, nearly all participants acknowledged the importance of family planning, with 97.5% of controls and 96.1% of cases expressing positive views. The most cited purpose of family planning was child spacing (52.1% of controls and 55% of cases), followed by benefits to maternal health, child health, and household economy.

Concerning knowledge of contraceptive types, 57.4% of controls and 44.7% of cases could identify methods suitable for both males and females. The average level of family planning knowledge was high for 76.6% of controls and 68% of cases. Ever use of family planning methods was similarly high in both groups (83.2% of controls and 83.5% of cases).

Among those who had used contraception, short-term methods were the most popular (59.1% of controls and 50% of cases), followed by long-term methods (37.8% of controls and 40.9% of cases), with natural methods being least utilized. The preference for contraceptive methods was primarily based on low side effects (24.4% of controls and 36% of cases, *p*=0.02) and accessibility.

The majority accessed family planning services free of charge (53.1% of controls and 54.9% of cases), while about 39% of both groups found services reasonably priced.

Among the reasons for not using contraception prior to the current pregnancy, children’s desire was cited by 68.8% of controls and 28.6% of cases. Fear of side effects was reported by 12.5% of controls compared to 42.9% of cases. Other reasons like religious beliefs, inaccessibility, and partner domination were less frequently reported.

At the time of the survey, 53.3% of respondents were in their third trimester, with 56.1% of controls and 50% of cases falling in this category. Regarding antenatal care (ANC) initiation, half of the participants began ANC during the first trimester, with no significant difference between groups.

Incorrect utilization of contraceptives was the leading reason for the current pregnancy among cases (37.9%), followed by contraceptive failure (27.4%) and partner preference (15.8%). Among controls, the most cited reason was partner preference (85.7%).

Among cases reporting contraceptive failure, the most frequently failed methods were the calendar method (30.8%), followed by IUD (19.2%), injectable (15.4%), and post-pill (15.4%).

Regarding parity, 41.3% of controls and 35% of cases were nulliparous. Primiparous women made up 31.6% of controls and 20% of cases, while multiparous women accounted for 27% of controls and 45% of cases, showing a statistically significant difference (*p*=0.02).

Ideal desired child number was three or more for 78.7% of controls and 58.3% of cases. Decision-making around pregnancy was predominantly joint in both groups, with 77.2% of controls and 60.1% of cases deciding together with their partners. However, decision-making by the partner alone was more common among cases (29.1%) than controls (11.7%, *p*=0.04) (<u>Table 2</u>).

**Table 2:**
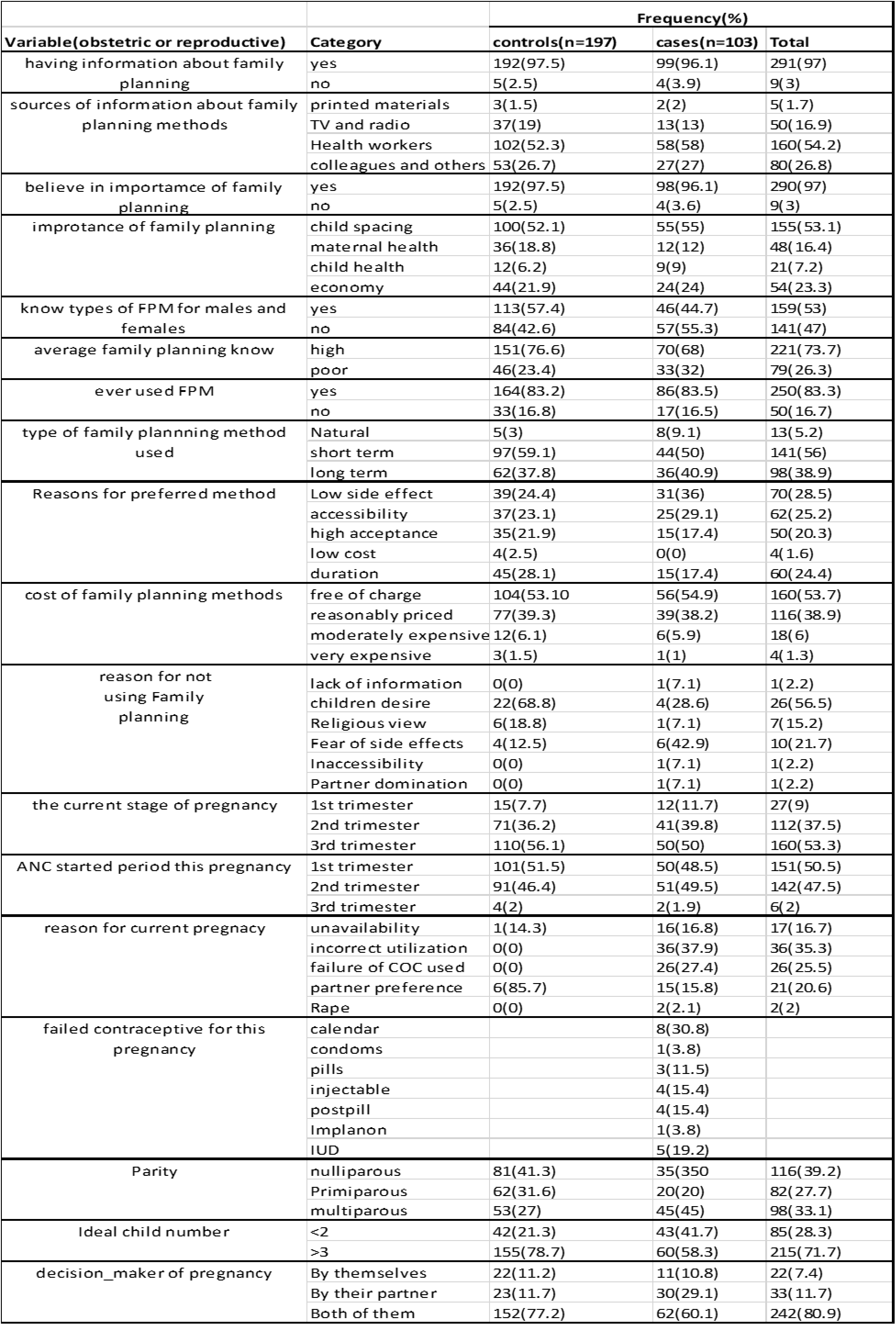
Obstetric and reproductive health related characteristics of unintended pregnancy, Lemi Kura sub-city, Addis Ababa, Ethiopia, 2022 (n=300).

| Variable(obstetric or reproductive) | Category | Frequency(%) |  |  |
| --- | --- | --- | --- | --- |
|  |  | controls(n=197) | cases(n=103) | Total |
| having information about family planning | yes | 192(97.5) | 99(96.1) | 291(97) |
|  | no | 5(2.5) | 4(3.9) | 9(3) |
| sources of information about family planning methods | printed materials | 3(1.5) | 2(2) | 5(1.7) |
|  | TV and radio | 37(19) | 13(13) | 50(16.9) |
|  | Health workers | 102(52.3) | 58(58) | 160(54.2) |
|  | colleagues and others | 53(26.7) | 27(27) | 80(26.8) |
| believe in importance of family planning | yes | 192(97.5) | 98(96.1) | 290(97) |
|  | no | 5(2.5) | 4(3.6) | 9(3) |
| importance of family planning | child spacing | 100(52.1) | 55(55) | 155(53.1) |
|  | maternal health | 36(18.8) | 12(12) | 48(16.4) |
|  | child health | 12(6.2) | 9(9) | 21(7.2) |
|  | economy | 44(21.9) | 24(24) | 54(23.3) |
| know types of FPM for males and females | yes | 113(57.4) | 46(44.7) | 159(53) |
|  | no | 84(42.6) | 57(55.3) | 141(47) |
| average family planning know | high | 151(76.6) | 70(68) | 221(73.7) |
|  | poor | 46(23.4) | 33(32) | 79(26.3) |
| ever used FPM | yes | 164(83.2) | 86(83.5) | 250(83.3) |
|  | no | 33(16.8) | 17(16.5) | 50(16.7) |
| type of family planning method used | Natural | 5(3) | 8(9.1) | 13(5.2) |
|  | short term | 97(59.1) | 44(50) | 141(56) |
|  | long term | 62(37.8) | 36(40.9) | 98(38.9) |
| Reasons for preferred method | Low side effect | 39(24.4) | 31(36) | 70(28.5) |
|  | accessibility | 37(23.1) | 25(29.1) | 62(25.2) |
|  | high acceptance | 35(21.9) | 15(17.4) | 50(20.3) |
|  | low cost | 4(2.5) | 0(0) | 4(1.6) |
|  | duration | 45(28.1) | 15(17.4) | 60(24.4) |
| cost of family planning methods | free of charge | 104(53.10) | 56(54.9) | 160(53.7) |
|  | reasonably priced | 77(39.3) | 39(38.2) | 116(38.9) |
|  | moderately expensive | 12(6.1) | 6(5.9) | 18(6) |
|  | very expensive | 3(1.5) | 1(1) | 4(1.3) |
| reason for not using Family planning | lack of information | 0(0) | 1(7.1) | 1(2.2) |
|  | children desire | 22(68.8) | 4(28.6) | 26(56.5) |
|  | Religious view | 6(18.8) | 1(7.1) | 7(15.2) |
|  | Fear of side effects | 4(12.5) | 6(42.9) | 10(21.7) |
|  | Inaccessibility | 0(0) | 1(7.1) | 1(2.2) |
|  | Partner domination | 0(0) | 1(7.1) | 1(2.2) |
| the current stage of pregnancy | 1st trimester | 15(7.7) | 12(11.7) | 27(9) |
|  | 2nd trimester | 71(36.2) | 41(39.8) | 112(37.5) |
|  | 3rd trimester | 110(56.1) | 50(50) | 160(53.3) |
| ANC started period this pregnancy | 1st trimester | 101(51.5) | 50(48.5) | 151(50.5) |
|  | 2nd trimester | 91(46.4) | 51(49.5) | 142(47.5) |
|  | 3rd trimester | 4(2) | 2(1.9) | 6(2) |
| reason for current pregnancy | unavailability | 1(14.3) | 16(16.8) | 17(16.7) |
|  | incorrect utilization | 0(0) | 36(37.9) | 36(35.3) |
|  | failure of COC used | 0(0) | 26(27.4) | 26(25.5) |
|  | partner preference | 6(85.7) | 15(15.8) | 21(20.6) |
|  | Rape | 0(0) | 2(2.1) | 2(2) |
| failed contraceptive for this pregnancy | calendar |  | 8(30.8) |  |
|  | condoms |  | 1(3.8) |  |
|  | pills |  | 3(11.5) |  |
|  | injectable |  | 4(15.4) |  |
|  | postpill |  | 4(15.4) |  |
|  | Implanon |  | 1(3.8) |  |
|  | IUD |  | 5(19.2) |  |
| Parity | nulliparous | 81(41.3) | 35(35) | 116(39.2) |
|  | Primiparous | 62(31.6) | 20(20) | 82(27.7) |
|  | multiparous | 53(27) | 45(45) | 98(33.1) |
| Ideal child number | <2 | 42(21.3) | 43(41.7) | 85(28.3) |
|  | >3 | 155(78.7) | 60(58.3) | 215(71.7) |
| decision_maker of pregnancy | By themselves | 22(11.2) | 11(10.8) | 22(7.4) |
|  | By their partner | 23(11.7) | 30(29.1) | 33(11.7) |
|  | Both of them | 152(77.2) | 62(60.1) | 242(80.9) |

Out of 250 (83.3%) participant practiced family planning method those used calendar were 5.2% (9.1% case and 3% control), condoms were 2% (2.3% case and 1.8% control), pills were 21% (19.3% case and 22% control), injectable were 32.9% (28.4% case and 35.4% control), Implanon were 29.8% (25% case and 32.3% control) and IUCD were 9.1% (15.9% case and 5.5%control)(Fig 2).

**Fig 2:**
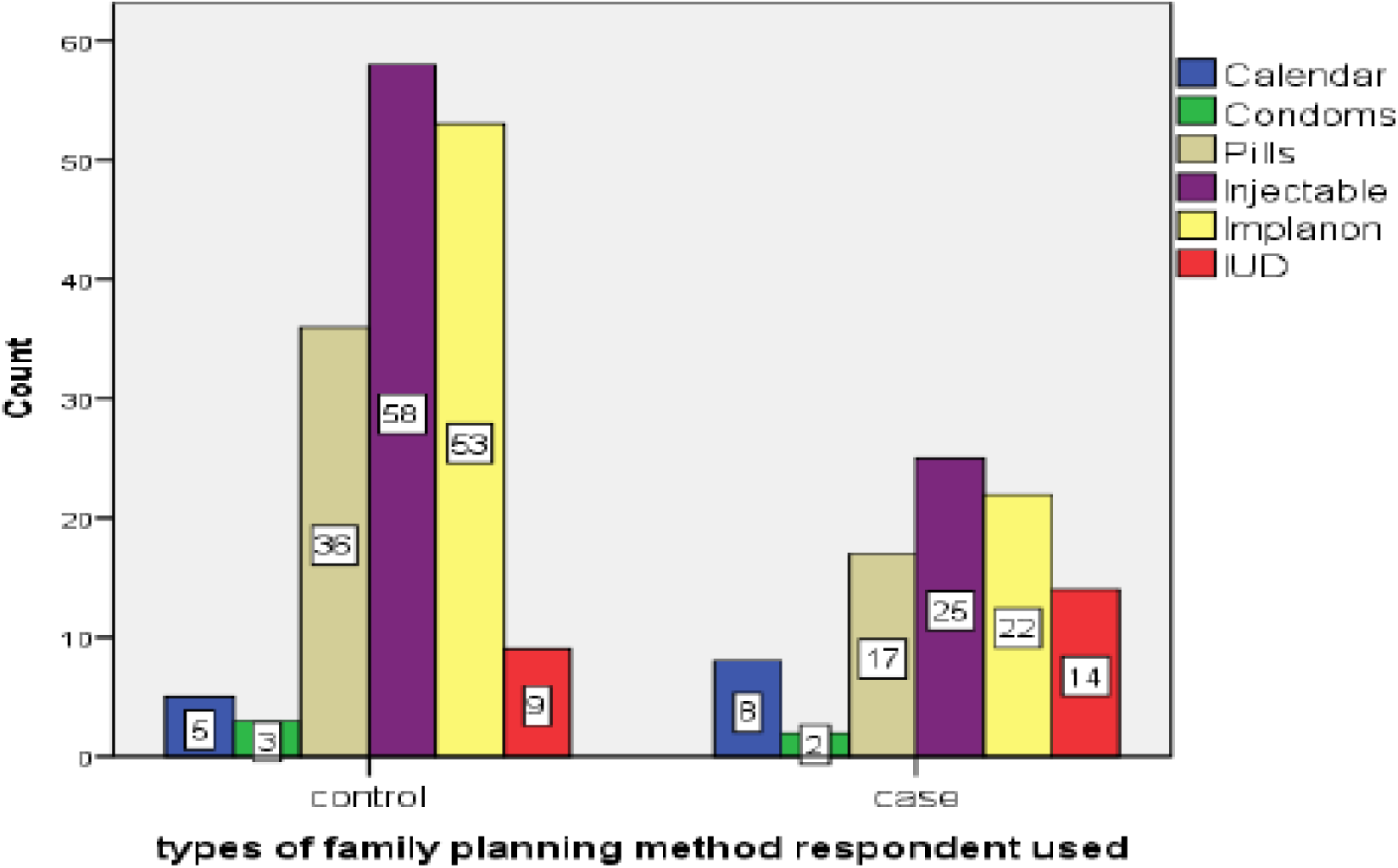
Types of family planning methods practiced among study participants Lemi Kura Sub city, Addis Ababa, Ethiopia. 2022 (n=300).

### 3.3 Behavioral characteristics of study participants

In this study, 10.7% of the participants reported drinking alcohol, with a higher proportion among cases (15.5%) compared to controls (8.2%). Khat chewing and cigarette smoking were each reported by 0.7% of participants, both slightly higher among cases (2.0%) than controls (1.0% and 0.5%, respectively). Additionally, having multiple sexual partners was more common among cases (23.3%) than controls (4.1%) <u>(Table 3).</u>

**Table 3:** Behavioral characteristics of unintended pregnancy, Lemi Kura sub-city, Addis Ababa, =Ethiopia, 2022 (n=300).

|  |  | Frequency(%) |  |  | P-value |
| --- | --- | --- | --- | --- | --- |
| behavioral characteristics | Category | controls(n=197) | cases(n=103) | Total |  |
| Alcohol | Drinking | 16(8.20) | 16(15.50) | 32(10.70) | 0.06 |
|  | Not drinking | 179(91.80) | 87(84.50) | 266(89.30) | 0.00 |
| khate | chewing | 2(1.00) | 0(0) | 2(0.70) |  |
|  | Not chewing | 195(99) | 102(100) | 297(99.30) |  |
| smoking | smoker | 1(0.50) | 1(1.00) | 2(0.70) |  |
|  | Not smoker | 196(99.50) | 101(99) | 297(99.30) |  |
| Multiple sexual partners | Have | 8(4.10) | 24(23.30) | 32(10.70) | 0.00 |
|  | Don't have | 189(95.90) | 79(76.70) | 268(89.30) | 0.00 |

### 3.4. Factors affecting the unintended pregnancy

Out of 16 variables identified through bivariate analysis for further investigation of multivariate analysis, only 7 variables were identified as determinants of unintended pregnancy.

Variables such as age of woman, marital status, dwelling status, religion, ethnicity, educational status, distance from an ANC service (a health facility), average knowledge about Family planning method, ever use of any type of family planning method, types of Family planning method they had used, reason for family planning method preference, parity, ideal number of children they need to have, the process how decision made about current pregnancy, experienced alcohol drinking, and having multiple sexual partners were identified as candidate determinants of unintended pregnancy during bivariate logistic regression analysis.

However, on multivariable logistic regression analysis, variables such as religion, distance from an ANC service (a health facility), and reason for family planning method preference, decision maker of pregnancy, parity, ideal child number, and having multiple sexual partners were identified as independent predictors of unintended pregnancy.

Muslim and Protestant women had 3.33 and 3.06 times the odds of experiencing an unintended pregnancy, respectively, compared to Orthodox Christian women (Muslim: AOR = 3.33; 95% CI: 1.21–9.15; p < 0.02; Protestant: AOR = 3.06; 95% CI: 1.07–8.75; p < 0.04).

Women who traveled 4 km or more from their home to the health facility for current ANC services had 2.70 times the odds of experiencing an unintended pregnancy compared to those who traveled less than 4 km (AOR = 2.70; 95% CI: 1.07–8.75; p < 0.02).

Compared to women who cited duration as their reason for preferring a family planning method, those who cited accessibility had 5.92 times the odds of experiencing an unintended pregnancy (AOR = 5.92; 95% CI: 1.41–24.79; p <0.02).

Multiparous women had 5.37 times the odds of experiencing an unintended pregnancy compared to nulliparous women (AOR = 5.37; 95% CI: 1.60–18.15; p < 0.01).

Women whose ideal number of children was less than or equal to two had 3.63 times the odds of experiencing an unintended pregnancy compared to women whose ideal number of children was three or more (AOR = 3.63; 95% CI: 1.50–9.01; p < 0.01).

Women whose current pregnancy decision was made by their partner had 16.58 times the odds of experiencing an unintended pregnancy compared to those who made the decision themselves (AOR = 16.58; 95% CI: 2.40–114.15; p < 0.01).

Women with multiple sexual partners had 6.46 times the odds of experiencing an unintended pregnancy compared to women with a single partner (AOR = 6.46; 95% CI: 1.38–30.38; p < 0.02) (Table 4).

**Table 4:**
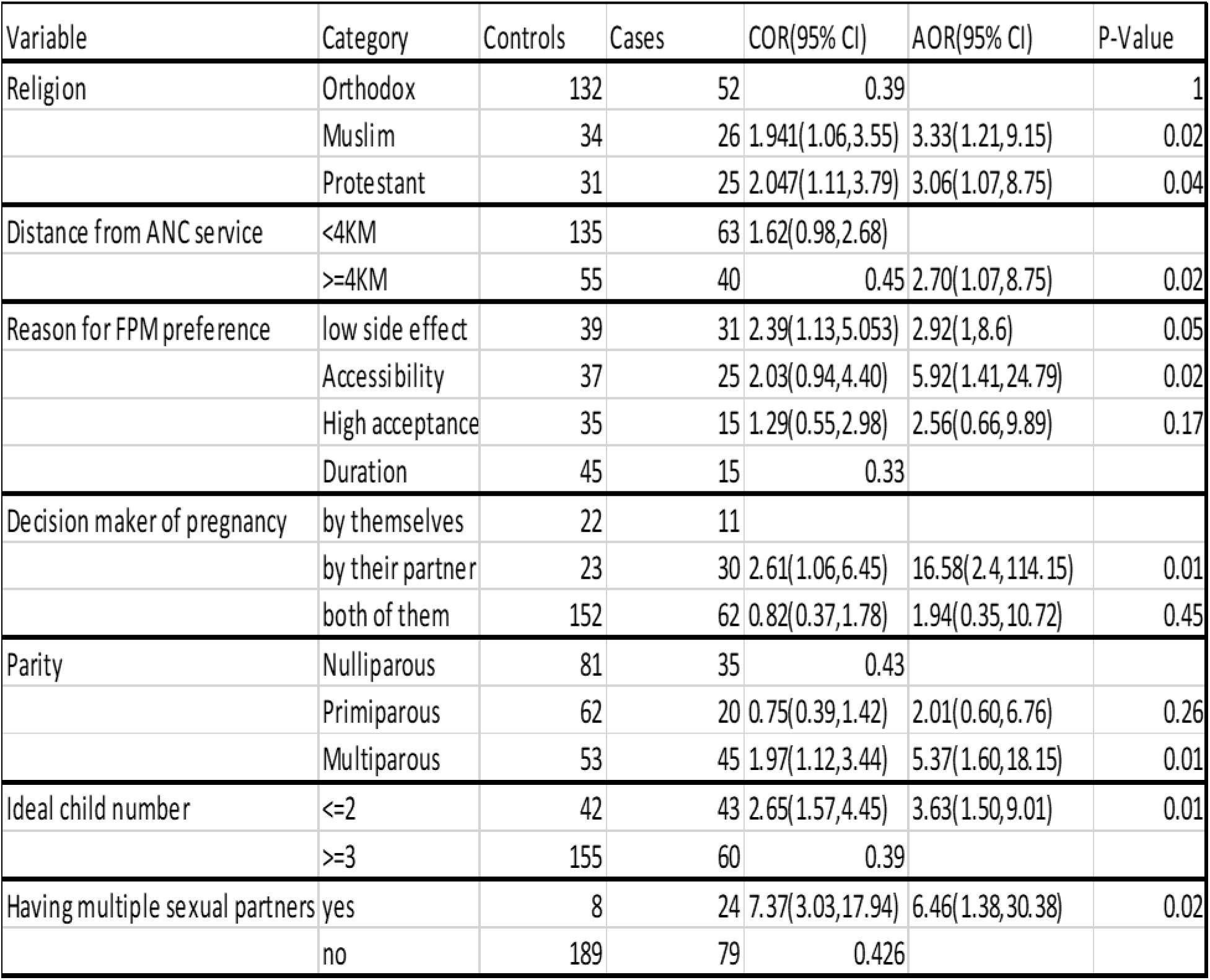
Factors associated with unintended pregnancy among pregnant women attending the ANC services at selected health care facilities, Lami Kura subcity, Addis Ababa, 2022.

## Discussion

The results of this study indicated that unintended pregnancy could occur as the result of socio-demographic factors like being Protestant or Muslim instead of Orthodox and visiting the health facilities somewhat far at least more than or equal to 4 km from their residence for ANC follow up. As the result of obstetric factors like dependent women instead of self-autonomous, being multiparous instead of nulli/ primiparous and when the Ideal number of children decreases. Finally, behavioral factors having multiple sexual partners. Therefore, in family planning programs, those women having two or more children and multiple sexual partners should be the focus of special attention by assuring the accessibility of contraceptives and creating clear awareness of the side effects of the programs to prevent unintended pregnancy and its consequences.

Muslim and Protestant pregnant women had 3.33 and 3.06 times higher odds of unintended pregnancy compared to Orthodox women, consistent with studies in India (<u>19</u>) (Muslim) and Wolayita, Ethiopia (<u>24</u>) (Protestant). This disparity may stem from lower autonomy and pronatalism in Muslim communities (<u>19</u>) and Protestant religious teachings discouraging contraceptive use (<u>24</u>). The observed differences from Ethiopian findings (<u>35</u>) could reflect variations in study design.

Pregnant women who traveled longer distances to ANC facilities had 2.70 times higher odds of unintended pregnancy compared to those visiting nearby facilities, aligning with studies in Ethiopia (<u>22</u>) and Kersa, eastern Ethiopia (<u>6</u>). This disparity may reflect lower service quality at local facilities and insufficient integration of family planning outreach programs with the Oromia Health Bureau in outlying areas like Finfine Zuria.

Multiparous women had 5.37 times higher odds of unintended pregnancy compared to nulliparous women, consistent with findings from Ecuador (<u>16</u>), Bangladesh (<u>17</u>), Iran (<u>9</u>), and Ethiopia (<u>22</u>). This likely reflects lower fertility desire among multiparous women due to accumulated gynecological, obstetric, and economic burdens.

Women desiring ≤2 children had 3.63 times higher odds of unintended pregnancy compared to those wanting ≥3 children, indicating an inverse relationship between ideal family size and unintended pregnancy. This aligns with findings from UNFPA (<u>1</u>), Nepal (<u>7</u>), and Ganji-Oromia, Ethiopia (<u>3</u>), potentially reflecting women’s limited autonomy in reproductive decision-making.

Women with low autonomy had 16.58 times higher odds of unintended pregnancy compared to autonomous women, consistent with findings from UNFPA (<u>1</u>), Bangladesh (<u>17</u>), Tanzania (<u>29</u>), and Ethiopia (<u>36</u>). This suggests enhancing women’s autonomy could address gender inequities while reducing unintended pregnancy and improving maternal-child health outcomes. The contrasting results from Nepal (<u>7</u>) may reflect methodological differences in autonomy measurement.

Women with multiple sexual partners had 6.46 times higher odds of unintended pregnancy than those with one partner, aligning with studies in China (<u>31</u>) and Tanzania (<u>33</u>). Potential explanations include inconsistent contraceptive use, alcohol-related risk behaviors, and reduced partner accountability in multi-partner relationships.

## Conclusions

This study identified several important predictors of unintended pregnancy, including partner-led pregnancy decisions, having multiple sexual partners, multiparity, a lower ideal number of children, religious affiliation, greater distance to health facilities for ANC services, and reasons for family planning method preference. These findings highlight the need for targeted interventions that promote women’s autonomy in reproductive decision-making, improve access to comprehensive family planning services, and address sociocultural and service accessibility barriers to reduce unintended pregnancies.

## Data availability statement

The authors confirm that the raw data supporting the findings of this study are available from the corresponding author upon reasonable request and without unnecessary restriction.

## Ethics statement

The study was conducted according to the Helsinki Declaration for Human Subjects Research (<u>37</u>). The Ethical approval was obtained from the Institutional Health Research Ethical Review Committee of the College of St. Paul Hospital Millennium Medical College (SPHMMC), Ethiopia (Ref.no: ሕ/ጤ/ት/716/2014 E.C). Written informed consent was secured from all participants after explaining the purpose and benefits of the study.

## Author contributions

AA: Conceptualization, Data curation, Formal analysis, Investigation, Methodology, Project administration, Resources, Software, Supervision, Validation, Visualization, writing – original draft, Writing – review & editing.

TT: Conceptualization, Data curation, Methodology, Software, Supervision, Validation, Visualization, writing – original draft, Writing – review & editing.

ST: Conceptualization, Data curation, Methodology, Software, Supervision, Validation, Visualization, writing – original draft.

KW: Conceptualization, Data curation, Methodology, Software, Validation, Visualization, writing – original draft, Writing – review & editing.

ND: Conceptualization, Data curation, Methodology, Software, Supervision, Validation, Visualization, writing – original draft, Writing – review & editing.

TA Conceptualization, Data curation, Methodology, Software, Supervision, Visualization, writing – original draft, Writing – review & editing.

AG: Conceptualization, Data curation, Methodology, Software, Supervision, Validation, Visualization, funding acquisition, writing – original draft, Writing – review & editing.

## Acknowledgements

First of all we thanks almighty God for his help, also authors would like to express their sincere gratitude to all the study participants, data collectors, and supervisors for their valuable contributions to this research. Special thanks are also extended to SPHMMC for providing financial support and to the staff of Lemi Kura health facilities for their cooperation and assistance in facilitating the data collection process.

